# Early clinical markers of aggressive multiple sclerosis

**DOI:** 10.1101/19002063

**Authors:** Charles B Malpas, Ali Manouchehrinia, Sifat Sharmin, Izanne Roos, Dana Horakova, Eva Kubala Havrdova, Maria Trojano, Guillermo Izquierdo, Sara Eichau, Roberto Bergamaschi, Patrizia Sola, Diana Ferraro, Alessandra Lugaresi, Alexandre Prat, Marc Girard, Pierre Duquette, Pierre Grammond, Francois Grand’Maison, Serkan Ozakbas, Vincent Van Pesch, Franco Granella, Raymond Hupperts, Eugenio Pucci, Cavit Boz, Gerardo Iuliano, Youssef Sidhom, Riadh Gouider, Daniele Spitaleri, Helmut Butzkueven, Aysun Soysal, Thor Petersen, Freek Verheul, Rana Karabudak, Recai Turkoglu, Cristina Ramo-Tello, Murat Terzi, Edgardo Cristiano, Mark Slee, Pamela McCombe, Richard Macdonell, Yara Fragoso, Javier Olascoaga, Ayse Altintas, Tomas Olsson, Jan Hillert, Tomas Kalincik, on behalf of the MSBase Study Group

**Author notes:** Corresponding author: Tomas Kalincik, Clinical Outcomes Research Unit (CORe), Department of Medicine, Royal Melbourne Hospital, The University of Melbourne, Melbourne, Australia. Disclosures of Interest: Charles Malpas: nothing to disclose. Sifat Sharmin: nothing to disclose. Dana Horakova received speaker honoraria and consulting fees from Biogen, Merck, Teva and Novartis, as well as support for research activities from Biogen and research grants from Charles University in Prague [PRVOUK-P26/LF1/4], Czech Minsitry of Education [PROGRES Q27/LF1] and Czech Ministry of Health [NT13237-4/2012]. Eva Kubala Havrdova received speaker honoraria and consultant fees from Actelion, Biogen, Celgene, Merck, Novartis, Roche, Sanofi and Teva, and support for research activities from Czech Ministry of Education [project Progres Q27/LF1]. Maria Trojano received speaker honoraria from Biogen-Idec, Bayer-Schering, Sanofi Aventis, Merck, Teva, Novartis and Almirall; has received research grants for her Institution from Biogen-Idec, Merck, and Novartis. Guillermo Izquierdo received speaking honoraria from Biogen, Novartis, Sanofi, Merck, Roche, Almirall and Teva. Sara Eichau received speaker honoraria and consultant fees from Biogen, Merck, Novartis, Almirall, Roche, Sanofi and Teva. Roberto Bergamaschi received speaker honoraria from Bayer Schering, Biogen, Genzyme, Merck, Novartis, Sanofi-Aventis, Teva; research grants from Bayer Schering, Biogen, Merck, Novartis, Sanofi-Aventis, Teva; congress and travel/accommodation expense compensations by Almirall, Bayer Schering, Biogen, Genzyme, Merck, Novartis, Sanofi-Aventis, Teva. Patrizia Sola served on scientific advisory boards for Biogen Idec and TEVA, she has received funding for travel and speaker honoraria from Biogen Idec, Merck, Teva, Sanofi Genzyme, Novartis and Bayer and research grants for her Institution from Bayer, Biogen, Merck, Novartis, Sanofi, Teva. Diana Ferraro received travel grants and/or speaker honoraria from Merck, TEVA,†Novartis, Biogen and Sanofi-Genzyme. Alessandra Lugaresi is a Bayer, Biogen, Genzyme, Merck Advisory Board Member. She received travel grants and honoraria from Roche, Bayer, Biogen, Merck, Novartis, Sanofi, Teva and Fondazione Italiana Sclerosi Multipla (FISM). Her institution received research grants from Bayer, Biogen, Merck, Novartis, Sanofi, Teva and Fondazione Italiana Sclerosi Multipla (FISM). Alexandre Prat did not declare any competing interests. Marc Girard received consulting fees from Teva Canada Innovation, Biogen, Novartis and Genzyme Sanofi; lecture payments from Teva Canada Innovation, Novartis and EMD.† He has also received a research grant from Canadian Institutes of Health Research. Pierre Duquette served on editorial boards and has been supported to attend meetings by EMD, Biogen, Novartis, Genzyme, and TEVA Neuroscience. He holds grants from the CIHR and the MS Society of Canada and has received funding for investigator-initiated trials from Biogen, Novartis, and Genzyme. Pierre Grammond is a Merck, Novartis, Teva-neuroscience, Biogen and Genzyme advisory board member, consultant for Merck, received payments for lectures by Merck, Teva-Neuroscience and Canadian Multiple sclerosis society, and received grants for travel from Teva-Neuroscience and Novartis. Francois Grand’Maison received honoraria or research funding from Biogen, Genzyme, Novartis, Teva Neurosciences, Mitsubishi and ONO Pharmaceuticals. Serkan Ozakbas did not declare any competing interests. Vincent Van Pesch received travel grants from Biogen, Bayer Schering, Genzyme, Merck, Teva and Novartis Pharma. His institution receives honoraria for consultancy and lectures from Biogen, Bayer Schering, Genzyme, Merck, Roche, Teva and Novartis Pharma as well as research grants from Novartis Pharma and Bayer Schering. Franco Granella received research grant from Biogen, served on scientific advisory boards for Biogen, Novartis, Merck, and Sanofi-Aventis and received funding for travel and speaker honoraria from Biogen, Merck, Sanofi-Aventis, and Almirall. Raymond Hupperts received honoraria as consultant on scientific advisory boards from Merck, Biogen, Genzyme-Sanofi and Teva, research funding from Merck and Biogen, and speaker honoraria from Sanofi-Genzyme and Novartis. Eugenio Pucci served on scientific advisory boards for Merck, Genzyme and Biogen; he has received honoraria and travel grants from Sanofi Aventis, Novartis, Biogen, Merck, Genzyme and Teva; he has received travel grants and equipment from “Associazione Marchigiana Sclerosi Multipla e altre malattie neurologiche”. Tomas Kalincik served on scientific advisory boards for Roche, Genzyme-Sanofi, Novartis, Merck and Biogen, steering committee for Brain Atrophy Initiative by Genzyme, received conference travel support and/or speaker honoraria from WebMD Global, Novartis, Biogen, Genzyme-Sanofi, Teva, BioCSL and Merck and received research support from Biogen. Cavit Boz received conference travel support from Biogen, Novartis, Bayer-Schering, Merck and Teva; has participated in clinical trials by Sanofi Aventis, Roche and Novartis. Gerardo Iuliano had travel/accommodations/meeting expenses funded by Bayer Schering, Biogen, Merck, Novartis, Sanofi Aventis, and Teva. Youssef Sidhom did not declare any competing interests. Riadh Gouider did not declare any competing interests. Daniele Spitaleri received honoraria as a consultant on scientific advisory boards by Bayer-Schering, Novartis and Sanofi-Aventis and compensation for travel from Novartis, Biogen, Sanofi Aventis, Teva and Merck. Helmut Butzkueven served on scientific advisory boards for Biogen, Novartis and Sanofi-Aventis and has received conference travel support from Novartis, Biogen and Sanofi Aventis. He serves on steering committees for trials conducted by Biogen and Novartis, and has received research support from Merck, Novartis and Biogen. Aysun Soysal did not declare any competing interests. Thor Petersen received funding or speaker honoraria from Biogen, Merck, Novartis, Bayer Schering, Sanofi-Aventis, Roche, and Genzyme. Freek Verheul is an advisory board member for Teva Biogen Merck and Novartis. Rana Karabudak did not declare any competing interests. Recai Turkoglu did not declare any competing interests. Cristina Ramo-Tello received research funding, compensation for travel or speaker honoraria from Biogen, Novartis, Genzyme and Almirall. Murat Terzi received travel grants from Novartis, Bayer-Schering, Merck and Teva; has participated in clinical trials by Sanofi Aventis, Roche and Novartis. Edgardo Cristiano received honoraria as consultant on scientific advisory boards by Biogen, Bayer-Schering, Merck, Genzyme and Novartis; has participated in clinical trials/other research projects by Merck, Roche and Novartis. Mark Slee has participated in, but not received honoraria for, advisory board activity for Biogen, Merck, Bayer Schering, Sanofi Aventis and Novartis. Pamela McCombe did not declare any competing. Richard Macdonell did not declare any competing interests. Yara Fragoso received honoraria as a consultant on scientific advisory boards by Novartis, Teva, Roche and Sanofi-Aventis and compensation for travel from Novartis, Biogen, Sanofi Aventis, Teva, Roche and Merck. Javier Olascoaga serves on scientific advisory boards for Biogen, Novartis, Sanofi and Roche; has received speaker honoraria from Almirall, Biogen, Bayer, Sanofi, Merck, Novartis and Roche and research grants from Biogen, Merck, Novartis and Teva. Ayse Altintas received personal fees and speaker honoraria from Teva, Merck, Novartis, Sanofi-Genzyme; received travel and registration grants from Merck, Roche, Sanofi-Genzyme. Tomas Olsson has received honoraria for lectures/advisory boards and/or unrestricted MS research grants from Biogen, Novartis, Sanofi, Merck and Roche. Data availability statement: Data were obtained from the international MSBase cohort study. Information regarding data availability can be obtained at https://www.msbase.org/.

## Abstract

Patients with the ‘aggressive’ form of MS accrue disability at an accelerated rate, typically reaching EDSS >= 6 within 10 years of symptom onset. Several clinicodemographic factors have been associated with aggressive MS, but less research has focused on clinical markers that are present in the first year of disease. The development of early predictive models of aggressive MS is essential to optimise treatment in this MS subtype. We evaluated whether patients who will develop aggressive MS can be identified based on early clinical markers, and to replicate this analysis in an independent cohort. Patient data were obtained from MSBase. Inclusion criteria were (a) first recorded disability score (EDSS) within 12 months of symptom onset, (b) at least 2 recorded EDSS scores, and (c) at least 10 years of observation time. Patients were classified as having ‘aggressive MS’ if they: (a) reached EDSS >= 6 within 10 years of symptom onset, (b) EDSS >=6 was confirmed and sustained over >=6 months, and (c) EDSS >=6 was sustained until the end of follow-up. Clinical predictors included patient variables (sex, age at onset, baseline EDSS, disease duration at first visit) and recorded relapses in the first 12 months since disease onset (count, pyramidal signs, bowel-bladder symptoms, cerebellar signs, incomplete relapse recovery, steroid administration, hospitalisation). Predictors were evaluated using Bayesian Model Averaging (BMA). Independent validation was performed using data from the Swedish MS Registry. Of the 2,403 patients identified, 145 were classified as having aggressive MS (6%). BMA identified three statistical predictors: age > 35 at symptom onset, EDSS >= 3 in the first year, and the presence of pyramidal signs in the first year. This model significantly predicted aggressive MS (AUC = .80, 95% CIs = .75, .84). The presence of all three signs was strongly predictive, with 32% of such patients meeting aggressive disease criteria. The absence of all three signs was associated with a 1.4% risk. Of the 556 eligible patients in the Swedish MS Registry cohort, 34 (6%) met criteria for aggressive MS. The combination of all three signs was also predictive in this cohort (AUC = .75, 95% CIs = .66, .84). Taken together, these findings suggest that older age at symptom onset, greater disability during the first year, and pyramidal signs in the first year are early indicators of aggressive MS.

---

Relapsing remitting multiple sclerosis (MS) is associated with the accrual of disability, typically due to incomplete recovery from relapses (Jokubaitis *et al*., 2016). A subset of patients accrue disability at an accelerated rate. While the rate of disability accrual in MS can represents a continuum, the European Committee for Treatment and Research in MS recognises that the rapidly progressing phenotype warrants a distinct clinical definition (European Committee for Treatment and Research in Multiple Sclerosis, 2018). An early attempt to define this group of patients was ‘malignant MS’, which was defined as “disease with a rapid progressive course, leading to significant disability in multiple neurologic systems or death in a relatively short time after disease onset” (Lublin and Reingold, 1996). It is estimated that malignant MS describes approximately 5-10% of patients (Gholipour *et al*., 2011).

More recently, the term ‘aggressive MS’ has gained increased acceptance (Gholipour *et al*., 2011). There is, however, no universally accepted definition of aggressive MS and a number of different criteria have been used. For example, in terms of the rate of disability accrual, Rush and colleagues suggest that reaching an expanded disability status scale (EDSS) of 4 within 5 years of symptom onset is a core feature of the phenotype (Rush *et al*., 2015). Some researchers used more severe criteria, requiring an EDSS of 6 or greater within 5 years (Menon *et al*., 2017), while others have required a gain of 2 or more EDSS points over a two year period (Scott *et al*., 2013). The diverse definitions used to describe this phenotype render interpretation of the literature challenging.

The determinants and clinical predictors of aggressive MS are not well understood; however a number of risk factors of more rapid disability accrual have been identified (see Rush *et al* (2015) for a recent review). Patient characteristics, such as being older at symptom onset or of male sex, have been recognised (Kantarci *et al*., 1998; Held *et al*., 2005; Tremlett *et al*., 2006; Ribbons *et al*., 2015). Several relapse characteristics have also been implicated, including: the presence of multifocal relapses (Kraft *et al*., 1981; Wolfson and Confavreux, 1987; Amato *et al*., 1999; Bergamaschi *et al*., 2001); partial or incomplete recovery from relapse; (Trojano *et al*., 1995; Scott and Schramke, 2010); greater frequency of relapses (Weinshenker *et al*., 1989; Confavreux *et al*., 2003; Ebers, 2005); and reduced time between relapses (Weinshenker *et al*., 1989; Trojano *et al*., 1995). Relapses associated with motor, cerebellar, bowel/balder, and cognitive signs have also been implicated (Citterio *et al*., 1989; Weinshenker *et al*., 1989; Phadke, 1990; Runmarker and Andersen, 1993; Weinshenker *et al*., 1996; Amato *et al*., 1999; Zarei *et al*., 2003; Langer-Gould *et al*., 2006; Kalincik, 2015). Neuroimaging signs have also been investigated, and the presence of high T2 lesion burden (Filippi *et al*., 1994; O’riordan *et al*., 1998; Brex *et al*., 2002; Rudick *et al*., 2006; Uher *et al*., 2017); multiple gadolinium-enhancing lesions (Kappos *et al*., 1999; Rudick *et al*., 2006); T1 lesions (Tomassini *et al*., 2006); early atrophy (Kornek and Lassmann, 2003; Lukas *et al*., 2010; Vaneckova *et al*., 2012); and infratentorial lesions (Sailer *et al*., 1999) have been shown to raise the risk of more rapid disability accrual.

While a number of risk factors have been described, the early clinical identification of patients at risk of aggressive MS presents a significant gap in current understanding. There is a paucity of large-scale observational studies that have interrogated multiple markers of aggressive MS simultaneously. To the best of our knowledge none have been conducted with the specific aim of clinical prediction and risk stratification. Critically, new evidence has demonstrated that early second-line therapy markedly improves disability outcomes over the long-term (Vaneckova *et al*., 2012; Fernández, 2017). However, identification of high-risk patients who are most likely to benefit from proactive treatment strategies remains an unmet need. The aim of this study was to define early clinical markers that identify patients who will later meet criteria for aggressive MS. To maximise clinical translation, we focused on markers are accessible to clinicians within the first 12 months following symptom onset. We focussed on estimating the risk of aggressive disease in individual patients and have conducted a validation in an independent cohort.

## Methods

### Participants

For the primary analysis, patient data were obtained from the international MSBase cohort study (WHO ICTRP ID: ACTRN12605000455662) (Butzkueven *et al*., 2006). The study was approved by the Melbourne Health Research Ethics Committee and site-specific ethics committees where relevant. All participants provided informed consent. Data from 58,589 patients attending 139 clinical centres in 34 countries were extracted from the MSBase registry in March 2018. Inclusion criteria were: (a) a diagnosis of clinically definite relapse-onset MS(Polman *et al*., 2011) (b) age at onset >= 18 years, (c) first expanded disability status score (EDSS) recorded within 12 months of symptom onset, (d) at least 2 recorded EDSS scores within 10 years of symptom onset, and (e) at least 10 years of observation time. All data were entered into the iMed patient record system or the MSBase online system as part of routine clinical practice. Data quality assurance procedures were performed as described elsewhere (Kalincik *et al*., 2017).

A second cohort of patients from the Swedish MS Registry (Hillert and Stawiarz, 2015) was used as an independent validation sample. Data from 19,520 patients were extracted from the registry in April 2018. The same inclusion criteria were applied as described above.

### Definition of aggressive MS

Patients were classified as having aggressive MS if they: (a) reached EDSS >= 6 within 10 years of symptom onset, (b) EDSS >=6 was confirmed and sustained over >=6 months, and (c) EDSS >=6 was sustained until the end of follow-up (>=10 years).

### Early clinical markers

Based on a survey of the relevant literature described above, a set of clinically translatable predictor variables was chosen. Patient characteristics included sex and age at symptom onset. The following predictors were included based on observation over the first 12 months following symptom onset: median EDSS, number of relapses, number of severe relapses (relapses that significantly affected activities of daily living or required hospitalisation), administration of steroids, hospitalisation (both used as a proxy for severe relapses), incomplete recovery after a relapse, pyramidal signs, bowel-bladder signs, cognitive signs, and cerebellar signs. The following nuisance variables were included: disease duration at first visit (maximum of 365 days), total observation time (minimum of 10 years), proportion of time spent on first-line (glatiramer acetate, interferon, dimethyl fumarate, teriflunomide) or second-line (mitoxantrone, natalizumab, cladribine, fingolimod, alemtuzumab, autologous stem cell transplantation, daclizumab, ocrelizumab, ofatumumab, rituximab, siponimod) therapy (Tramacere *et al*., 2015) over the first year since symptom onset, and proportion of time spent on first and second line therapy over the first 10 years since symptom onset. Magnetic resonance imaging (MRI) findings in the first year since symptom onset (presence of gadolinium enhancing, infratentorial, juxtatentorial, or periventricular lesions) and oligoclonal band results (positive or negative) were available for a subset of patients.

### Statistical analyses

All statistical analyses were performed in *R* (Version 3.4.4) (R Core Team, 2019). Generalised linear models (GLMs) were used to determine the statistical predictors of aggressive disease status. Aggressive disease was entered as the binary dependent variable and a binomial response distribution with logistic link function was chosen. The following baseline variables were entered as predictors: gender, age at symptom onset. The following events within the first year of symptom onset were also entered: median EDSS, any hospitalisation associated with a relapse, any treatment with steroids, number of severe relapses, number of any relapses, pyramidal signs, bowel/bladder signs, cerebellar signs, partial recovery from a relapse. The following nuisance variables were also entered: disease duration at first visit, disease duration at last visit (total observation time), proportion of time over the first year on first or second-line therapy, proportion of time over the 10-year observation period on first or second-line therapy.

The use of these 17 predictors of interest resulted in 2^17^ = 131,072 possible statistical models (i.e., different combinations of predictors). The selection of variables represents a well understood source of bias and it is difficult to know whether a particular set of predictors represents the ‘true’ model (Miller, 2002; Lukacs *et al*., 2010). We used a Bayesian model averaging (BMA) to address this issue (Raftery *et al*., 1997). Briefly, for *p* predictors BMA estimates all possible models corresponding to the 2^*p*^ possible combinations of predictors. The fit of each model was then evaluated using the log of the posterior odds. Parameter estimates were averaged over all models, weighted for the fit of each model, which inherently adjusted for the uncertainty associated with model selection bias.

The importance of each predictor was evaluated by the posterior inclusion probability (PIP) value, which was the probability that the parameter is not zero given the data. More formally, this is given as *P*(*B*≠ 0|*D*) where *B* is the parameter estimate for the predictor and *D* are the data. This value was computed from the posterior distribution of *B*, which is given as:

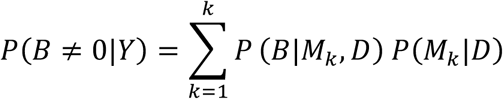

Where *B* is the parameter estimate for the predictor, *D* are the data, and *M1* to *Mk* are the models considered. This therefore represented the average of the posterior distribution for each parameter averaged over all models and weighted by the posterior probability (Hoeting *et al*., 1999). Predictors with PIP values > .5 were considered ‘important’. Predictions of aggressive disease status for each patient were computed using the posterior predictive distribution averaged across all models. Two sensitivity analyses were conducted. First, Bayesian generalised linear mixed models were computed to investigate the importance of modelling a random intercept across clinical sites. Second, all BMA analyses were repeated using a subgroup of patients who had an EDSS < 6 at first visit.

Receiver operating characteristic (ROC) curves were then computed for the combined model, the ‘reduced’ model containing important predictors only (also known as the median probability model), and for each of the important continuous predictors separately. The optimal cut-off probability was determined using Youden’s method (Youden, 1950). Key analyses were repeated using conventional frequentist generalised linear models, which were estimated using iteratively reweighted least squares. Robust standard errors were computed via heteroskedasticity-consistent sandwich estimation. Confidence intervals were computed based on these standard errors and are presented at the 95% level. Robust *p* values were also computed, with the critical α set at .05.

BMA was performed using the Bayesian adaptive sampling package (Clyde *et al*., 2011). The intercept was included in all models. The robust mixture of g-priors defined by Bayarri and colleagues (2012) was placed on each model coefficient. A beta-binomial prior was placed on each model with parameters α = 1 and β = 1. Bayesian adaptive sampling without replacement was used with uniform sampling probabilities specified across all parameters (Clyde *et al*., 2011). Bayesian generalised linear mixed models were estimated using the *brms* package to evaluate the need for random terms nested within clinical site (Bürkner, 2017, 2018). ROC curves were computed using the *pROC* package (Robin *et al*., 2011). Frequentist robust standard errors were computed using the *sandwich* package (Zeileis, 2004, 2006). The best model identified by BMA was replicated in the Swedish MS Registry cohort. Only the clinical predictors most strongly supported by the evidence were used for replication, with beta coefficients taken from the original model (rather than being re-estimated).

## Results

### Sample characteristics

The included sample comprised 2,403 patients, of whom 145 (6%) met criteria for aggressive disease. The average time from symptom onset to meeting aggressive disease criteria was 6.05 years (SD = 2.79, range = 0 – 9.89 years). As shown in *Table 1*, patients with aggressive disease tended to be older at symptom onset, have higher median EDSS during the first year, and a greater number of relapses in the first year. At a descriptive level of analysis, they were also more likely to experience hospitalisation, partial recovery from relapse, cerebellar signs, bowel/bladder signs, and pyramidal signs within the first year of symptom onset.

**Table 1.** Sample characteristics

| *Variable* | *Total*<br>*(N = 2,403)* | *Non-aggressive disease*<br>*(n = 2,258)* | *Aggressive disease*<br>*(n = 145)* |
| --- | --- | --- | --- |
| *Baseline variables* |  |  |  |
| Age at symptom onset | 31.72 (8.91) | 31.3 (8.61) | 38.4 (10.8) |
| Female – n (%) | 1712 (71.2) | 1613 (71.43) | 99 (68.28) |
| Disease duration at first visit (days) | 119.90 (110.0) | 118.00 (110.00) | 143.00 (113.00) |
| First year EDSS | 1.78 (1.26) | 1.69 (1.16) | 3.11 (1.83) |
| *Events within first 12 months* |  |  |  |
| No. relapses | 0.74 (0.93) | 0.72 (0.92) | 1.06 (0.96) |
| No. severe relapses | 0.12 (0.40) | 0.20 (0.40) | 0.17 (0.44) |
| Treatment with steroids – n (%) | 491 (20.4) | 448 (19.84) | 43 (29.66) |
| Hospitalisation – n (%) | 288 (12.0) | 257 (11.38) | 31 (21.38) |
| Partial recovery from relapse – n (%) | 186 (7.7) | 164 (7.26) | 22 (15.17) |
| Pyramidal signs – n (%) | 795 (33.1) | 699 (30.96) | 96 (66.21) |
| Bowel/bladder signs – n (%) | 154 (6.4) | 125 (5.54) | 29 (20) |
| Cerebellar signs – n (%) | 388 (16.1) | 332 (14.70) | 56 (38.62) |
| *Treatment* |  |  |  |
| % time on first-line (10y) | 46.00 (36.10) | 46.30 (36.50) | 40.80 (29.99) |
| % time on second-line (10y) | 5.30 (14.1) | 4.90 (13.7) | 11.30 (18.2) |
| % time on first-line (1y) | 17.1 (28.00) | 17.2 (28.2) | 14.80 (23.4) |
| % time on second-line (1y) | 0.50 (5.1) | 0.40 (4.80) | 1.40 (7.70) |

**Table 2.** BMA analysis for prediction of aggressive disease status

| *Variable* | *B* | *SD* | *95% CI* | *Exp(B)* | *PIP* |
| --- | --- | --- | --- | --- | --- |
| *Baseline variables* |  |  |  |  |  |
| Age at symptom onset | 0.07 | 0.01 | 0.05, 0.09 | 1.07 | **1.00** |
| Male | 0.01 | 0.06 | 0.00, 0.00 | 1.01 | 0.06 |
| Disease duration at first visit | 0.00 | 0.00 | 0.00, 0.00 | 1.00 | 0.15 |
| First year EDSS | 0.50 | 0.07 | 0.36, 0.64 | 1.65 | **1.00** |
| *Events within first 12 months* |  |  |  |  |  |
| No. relapses | 0.04 | 0.10 | 0.00, 0.29 | 1.04 | 0.25 |
| No. severe relapses | -0.01 | 0.05 | 0.00, 0.00 | 0.99 | 0.06 |
| Treatment with steroids | 0.01 | 0.07 | 0.00, 0.00 | 1.03 | 0.06 |
| Hospitalisation | 0.02 | 0.12 | 0.00, 0.20 | 1.03 | 0.09 |
| Partial recovery from relapse | 0.00 | 0.05 | 0.00, 0.00 | 1.00 | 0.04 |
| Pyramidal signs | 0.47 | 0.36 | 0.00, 1.00 | 1.60 | **0.75** |
| Bowel/bladder signs | 0.02 | 0.12 | 0.00, 0.14 | 1.02 | 0.09 |
| Cerebellar signs | 0.07 | 0.19 | 0.00, 0.59 | 1.07 | 0.20 |
| *Treatment* |  |  |  |  |  |
| % time on first-line (10y) | 0.00 | 0.05 | 0.00, 0.00 | 1.00 | 0.04 |
| % time on second-line (10y) | 2.32 | 0.52 | 1.37, 3.36 | 10.15 | **1.00** |
| % time on first-line (1y) | -0.01 | 0.10 | 0.00, 0.00 | 0.99 | 0.06 |
| % time on second-line (1y) | -0.05 | 0.37 | 0.00, 0.00 | 0.95 | 0.06 |

**Note:** *B* = model coefficients computed as the mean of the posterior distribution. SE = standard deviation of the posterior distributions. CI = 95% credible intervals. Exp(*B*) = odds ratios of the model coefficients. $P(B \neq 0 | Y)$ = posterior inclusion probability (PIP).

### Predictors of aggressive disease

BMA revealed several important predictors of aggressive disease. The top 20 models, sorted by posterior probability, are showing in *Figure 1*. The posterior inclusion probabilities (PIPs) are shown in *Figure 2*. The strongest evidence was observed for age at symptom onset and first year EDSS (PIPs = 1). The presence of pyramidal signs was also strongly supported (PIP = 0.75), as was the proportion of time spent on second-line treatment over 10 years (PIP = 1.00). As shown in *Table* 2, the remaining predictors had PIPs < .5. When averaged across the whole model space, the standardised parameter estimates for age of symptom onset, first year EDSS, and time spent on second-line therapy were significant independent predictors of aggressive disease, with their respective confidence intervals not capturing zero. The estimate for pyramidal signs, however, was less precise and included 0 as a plausible value at the 95% level. Taken in the context of the PIP value greater than .5, this indicates that this predictor is important to include in any well-fitting model, but there is considerable uncertainty regarding the specific magnitude of this relationship.

**Figure 1.**
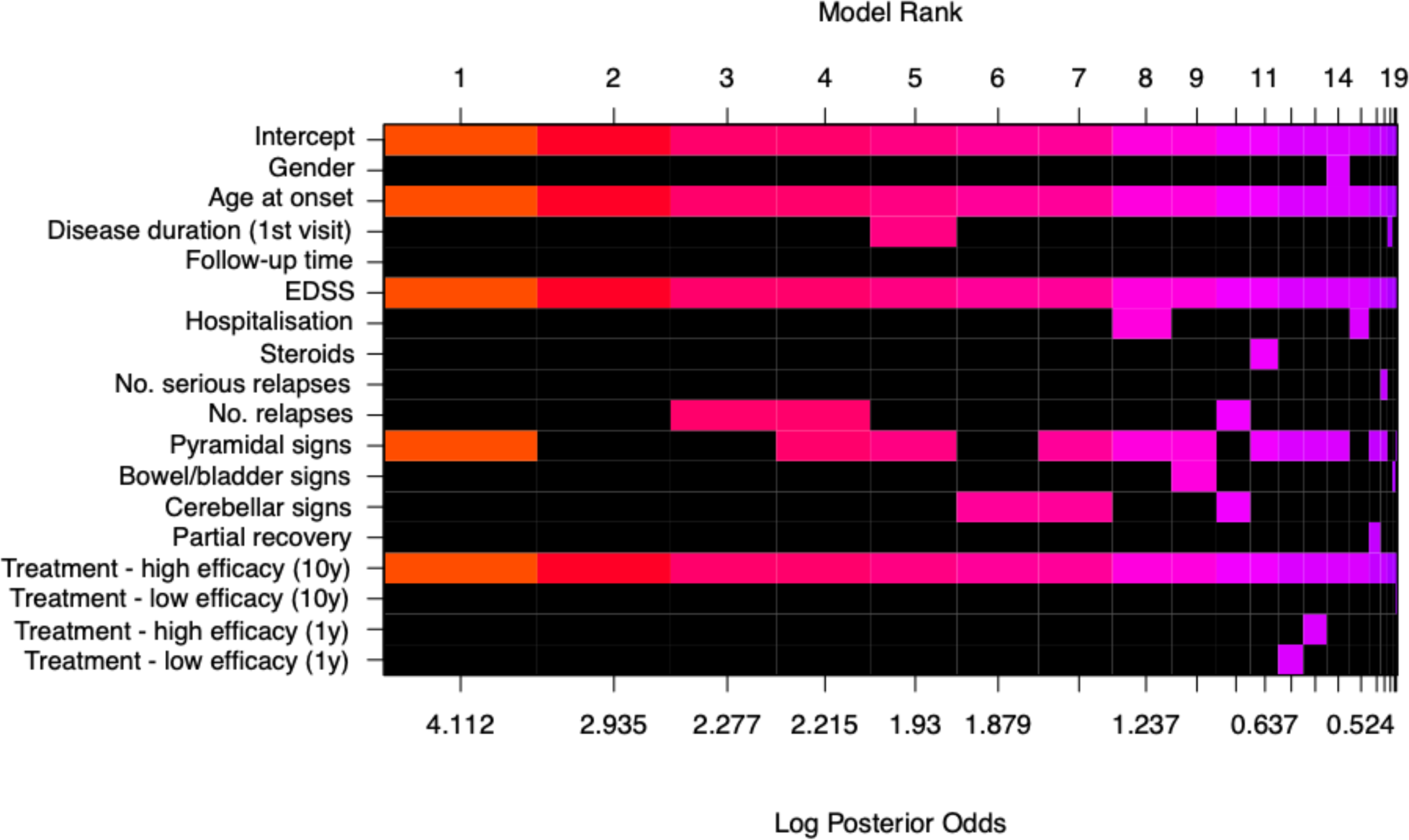
Visualisation of the model space from the Bayesian model averaging (BMA) analysis. The top 20 model are shown ranked by the log of the posterior odds. The colour indicates the posterior probability of the individual predictors (warmer = higher probability). Black squares indicate that the predictor was not included in the model. As shown, age at onset, EDSS, and pyramidal signs were consistently included in the best fitting models. The intercept was included in all models.

**Figure 2.**
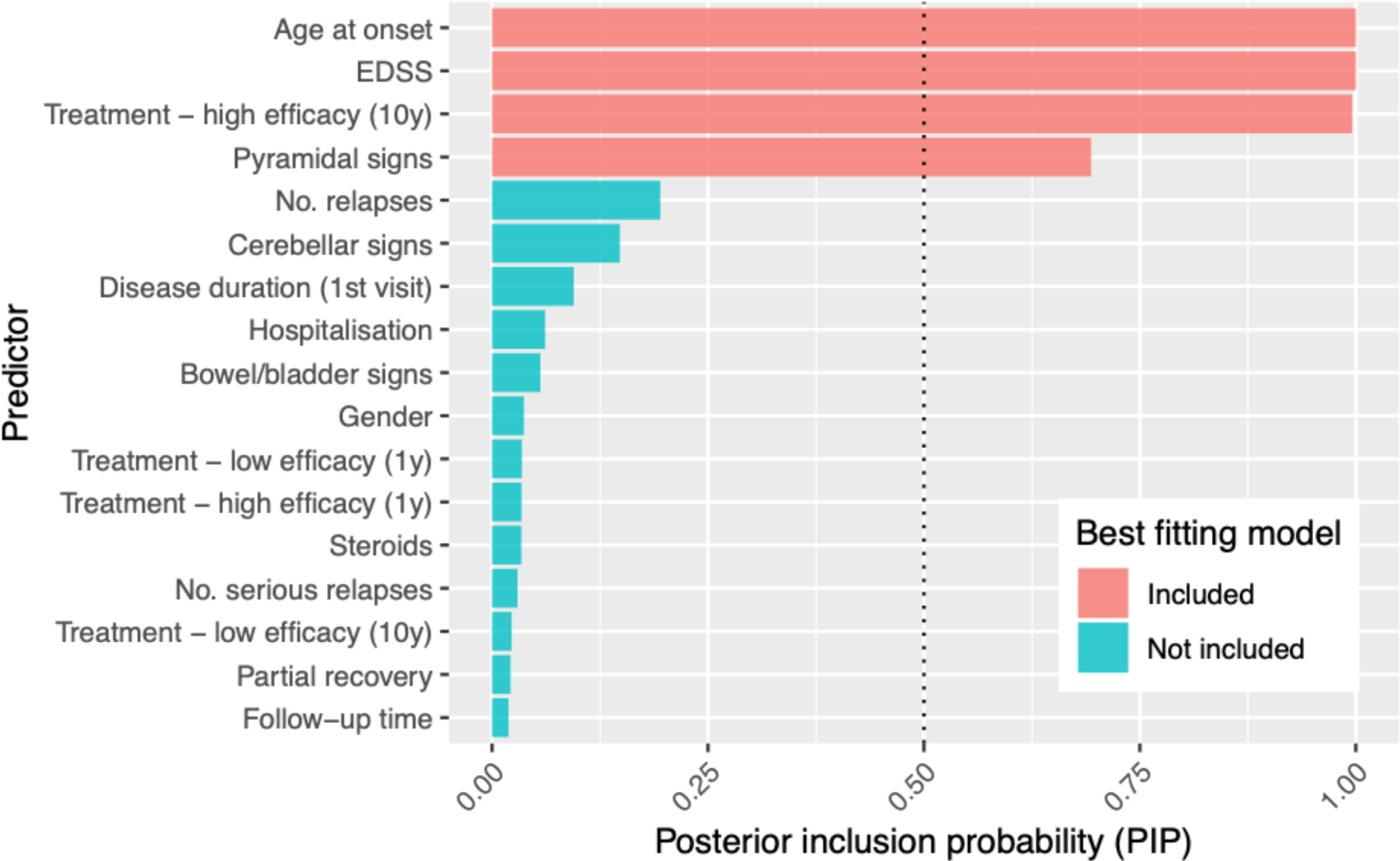
Posterior inclusion probabilities (PIP) for each predictor and model parameters averaged across the model space. EDSS, age at symptom onset, presence of pyramidal signs during the first year since symptom onset, and time spent on first-line therapy over 10 years were important predictors (PIPs > .5). All of these predictors were included in the single best fitting model. The evidence was not compelling for the other predictors.

### MRI and OCB

The same Bayesian model averaging approach was repeated in the subgroup of patients with information about cerebral MRI (*n = 359*) and CSF oligoclonal bands (*n* = 1,169) available. For the MRI data, the presence of gadolinium enhancing, infratentorial, juxtatentorial, or periventricular lesions were added to the full set of predictors as described above. For the OCB data, positive OCB status was added to the full set of predictors. All MRI markers had PIPs < .5 (range .01 – .04) which suggested they were not important predictors in the model. The PIP for oligoclonal bands positive status was .07, which also indicates that this was not an important predictor of aggressive disease. Model output for these analyses is included in *Appendix A*.

### Sensitivity analyses and assumption checks

The first sensitivity analysis was performed to investigate whether the clustering of cases within clinical sites should be explicitly modelled via a random effects analysis. Two Bayesian generalised linear models were estimated; the first with the intercept fixed across all cases, and the second with a random intercept for each clinical site. The fit of each model was computed in the form of the leave-one out information criteria (LOOIC). As described in *Appendix B*, the fixed intercept model was a better fit to the data (LOOIC = 922.19) compared to the random intercept (LOOIC = 923.20). As such, the fixed intercept model, as implemented in the BMA analysis, was deemed most appropriate for all analyses.

A second sensitivity analysis was performed in the subgroup of patients who had an EDSS < 6 at the time of first visit. As shown in *Appendix C*, these results largely replicated the findings of the main analysis. Age at symptom onset (PIP = 1.00), EDSS at first visit (PIP = 1.00), and the presence of pyramidal signs in the first year (PIP = .74) were all important predictors. The proportion of time spend on a second-line therapy over the 10-year observation period was also an important predictor (PIP = .99).

### Frequentist analysis

All analyses were repeated using a conventional frequentist approach. As shown in *Table 3*, the strongest evidence supported age and symptom onset and first year EDSS as significant predictors of aggressive MS (*p* < .001). Relatively weaker evidence was observed for the presence of pyramidal signs (*p* = .017). The proportion of time spent on second-line treatment over 10 years was statistically significance (*p* < .001). As shown in *Appendix C*, these findings were confirmed in the subgroup of patients with EDSS < 6 at first clinical visit.

**Table 3.** Frequentist GLM analysis for prediction of aggressive disease status

| *Variable* | *B* | *SE* | *95% CIs* | *Exp(B)* | *Sig.* |
| --- | --- | --- | --- | --- | --- |
| *Baseline variables* |  |  |  |  |  |
| Age at symptom onset | 0.07 | 0.01 | 0.05, 0.09 | 1.07 | **<0.001** |
| Male | 0.26 | 0.21 | -0.15, 0.66 | 1.29 | 0.210 |
| Disease duration at first visit | 0.00 | 0.00 | 0.00, 0.00 | 1.00 | 0.223 |
| First year EDSS | 0.43 | 0.09 | 0.26, 0.60 | 1.54 | **<0.001** |
| *Events within first 12 months* |  |  |  |  |  |
| No. relapses | 0.18 | 0.12 | -0.05, 0.41 | 1.20 | 0.122 |
| No. severe relapses | -0.33 | 0.23 | -0.78, 0.12 | 0.72 | 0.147 |
| Treatment with steroids | 0.08 | 0.26 | -0.42, 0.59 | 1.09 | 0.745 |
| Hospitalisation | 0.25 | 0.32 | -0.37, 0.88 | 1.29 | 0.429 |
| Partial recovery from relapse | 0.03 | 0.32 | -0.59, 0.65 | 1.03 | 0.935 |
| Pyramidal signs | 0.56 | 0.24 | 0.10, 1.03 | 1.76 | **0.017** |
| Bowel/bladder signs | 0.32 | 0.29 | -0.26, 0.89 | 1.38 | 0.276 |
| Cerebellar signs | 0.34 | 0.23 | -0.10, 0.78 | 1.41 | 0.130 |
| *Treatment* |  |  |  |  |  |
| % time on first-line (10y) | -0.19 | 0.32 | -0.80, 0.43 | 0.83 | 0.554 |
| % time on second-line (10y) | 2.27 | 0.58 | 1.14, 3.41 | 9.71 | **<0.001** |
| % time on first-line (1y) | -0.29 | 0.43 | -1.14, 0.55 | 0.75 | 0.497 |
| % time on second-line (1y) | -1.36 | 1.63 | -4.56, 1.84 | 0.26 | 0.404 |

**Note:** *B* = model coefficients. *SE* = robust standard errors computed via heteroskedasticity-consistent sandwich estimation. *CI* = 95% confidence intervals computed using robust *SEs*. *Exp(B)* = odds ratios for the model coefficients. *Sig* = significance values were computed using the robust *SEs*. Bold significance values indicate $p < .05$ . Assumption checks regarding multicollinearity, influential cases, autocorrelation, dispersion, and residual distribution were satisfied.

### Individual predictive accuracy

When averaged across the model space using BMA, the overall model was predictive of aggressive disease status. The area under the curve was .82 [95% CI = .78, .85] for the overall model, and .80 [95% CI = .75, .84] when using only the three important predictors (reduced model). Using the optimal cut-off of 0.06, the positive predictive value was 15%, compared to sample prevalence of aggressive disease of 6%. The negative predictive value was 98%. As shown in *Table 4*, ROC curves were also computed for each of the numerical predictors separately (age at symptom onset and median EDSS during the first year). Optimal thresholds were also computed, with patients classified as positive for each sign if their value was greater than the respective threshold (except for pyramidal signs, which were already binary). This procedure was repeated for a model containing all three of the important predictors identified above (age at symptom onset, median EDSS during the first year, pyramidal signs).

**Table 4.** Classification performance of the BMA analysis and specific variables

| *Variable* | *AUC*<br>*[95% CIs]* | *Threshold* | *Sensitivity* | *Specificity* | *PPV* | *NPV* |
| --- | --- | --- | --- | --- | --- | --- |
| BMA (full) | .82 [.78, .85] | .05 | .78 | .71 | .15 | .98 |
| BMA (reduced) | .80 [.75, .84] | .06 | .72 | .73 | .15 | .98 |
| Age at onset | .69 [.64, .74] | 35 | .60 | .71 | .12 | .96 |
| First year EDSS | .74 [.70, .79] | 2.5 | .53 | .85 | .19 | .97 |
| Pyramidal signs | - | - | .66 | .69 | .12 | .97 |

**Note:** AUC = area under the curve. Threshold = the optimal cut-off determined using Youden's method (1955). PPV = positive predictive value. NPV = negative predictive value. As the presence pyramidal signs is a binary predictor, the AUC and threshold could not be computed. BMA (full) = the entire model with all 17 predictors. BMA (reduced) = the model with only age at onset, first year EDSS, and pyramidal signs.

The number of positive signs was then translated into the probability of developing aggressive disease. Of the patients with all three signs, 32% had aggressive disease. This was followed by any two signs (14%), any one sign (5%), and no signs (1%). The specific combinations of positive signs were also computed. As shown in *Table 5*, this revealed a more complex pattern of relationships. For example, being positive for EDSS and pyramidal signs was associated with slightly lower risk of aggressive disease (11%) compared to being positive for age and EDSS (21%).

**Table 5.** Prognostic value of combinations of positive signs

| *Age*<br>*> 35* | *EDSS*<br>*>= 3* | *Pyramidal*<br>*signs* | *n* | *Non-aggressive*<br>*disease/aggressive*<br>*disease - n* | *% aggressive*<br>*disease* | *Relative*<br>*risk (RR)* |
| --- | --- | --- | --- | --- | --- | --- |
| ✓ | ✓ | ✓ | 125 | 85/40 | 32.00 | 22.85 |
| ✓ | ✓ |  | 52 | 41/11 | 21.15 | 15.11 |
| ✓ |  | ✓ | 177 | 154/23 | 12.99 | 9.28 |
|  | ✓ |  | 88 | 78/10 | 11.36 | 8.11 |
|  | ✓ | ✓ | 142 | 126/16 | 11.27 | 8.05 |
|  |  | ✓ | 351 | 334/17 | 4.84 | 3.46 |
| ✓ |  |  | 397 | 384/13 | 3.27 | 2.34 |
|  |  |  | 1071 | 1056/15 | 1.40 | *Ref.* |

### External validity

The performance of the three important predictors was evaluated in an independent sample obtained from the Swedish MS Registry. The mean age at symptom onset (*M* = 33.35, *SD* = 8.85) and median EDSS during the first year (*M* = 1.51, *SD* = 1.28) were comparable to the MSBase cohort. The proportion of patients reported to show motor signs in the first year (*n* = 95, 17%) was lower than the proportion of MSBase patients with pyramidal signs in the first year (*n* = 795, 33%). Of the 556 patients, 34 met criteria for aggressive disease (6%). This prevalence was comparable to the MSBase cohort. The probability of meeting aggressive disease criteria was predicted in the Swedish MS Registry using three important predictors identified in the MSBase cohort only. The area under the curve was .75 [95% CI .66, .84], only marginally lower than in the MSBase cohort, and within the confidence limits. The ROC is visualised in *Appendix D*. Using the optimal cut-off of 0.06 (determined from the primary analysis in the MSBase cohort), the positive predictive value was 15%, compared to sample prevalence of 6%. The negative predictive value was 97%.

## Discussion

In this study, we investigated a comprehensive set of early predictors of aggressive MS. Using data from the international MSBase registry (Butzkueven *et al*., 2006), we identified early and clinically accessible makers that, if observed in the first year since symptom onset, convey increased risk of the patient meeting criteria for aggressive disease. Specifically, older age at symptom onset, greater disability in the first year, and the presence of pyramidal signs are associated with aggressive MS. Conversely, the absence of these signs conveyed lower probability of meeting aggressive MS criteria compared to the observed base rate. The predictive power of the overall model was replicated using an independent cohort of patients from the Swedish MS registry. The Bayesian analytical approach enabled us to estimate posterior probabilities of aggressive disease which can be directly applied to individual patients who have recently presented with symptoms of relapse-onset MS.

Overall, these findings are consistent with a number of previous studies. Several studies have reported general associations of older age at symptom onset (Confavreux *et al*., 2003; Held *et al*., 2005; Confavreux and Vukusic, 2006), early disability progression (Weinshenker *et al*., 1989), and motor signs with accelerated worsening of disability (Weinshenker *et al*., 1989; Atkins and Freedman, 2013; Correale *et al*., 2014). In a number of areas, our results may seem to diverge from previously reported findings. For example, we did not find evidence for an independent, unequivocal effect of sex (Kantarci *et al*., 1998; Tremlett *et al*., 2006), partial recovery from relapse (Trojano *et al*., 1995; Scott and Schramke, 2010), severity of relapse (Rush *et al*., 2015), the number of relapses early in the disease course (Weinshenker *et al*., 1989; Confavreux *et al*., 2003; Ebers, 2005), and relapses presenting with cerebellar (Weinshenker *et al*., 1989; Phadke, 1990; Amato *et al*., 1999) or bowel/bladder (Citterio *et al*., 1989; Runmarker and Andersen, 1993; Amato *et al*., 1999; Langer-Gould *et al*., 2006) signs on the risk of developing aggressive disease. Due to issues of multi-collinearity, we were unable to investigate the independent effect of cognitive impairment, which has previously been suggested to be a poor prognostic sign (Zarei *et al*., 2003). The main conceptual difference between the present study and the previously published work is its focus on an objectively defined aggressive disease form rather than the overall risk of disability worsening used as the outcome in this study. Therefore, it is not surprising that several markers, which are predictive of disability worsening in general, are not directly associated with the risk of developing aggressive disease – a disease form that is at an extreme of the continuum of disease severity.

Identifying patients who are at a high risk of developing an aggressive MS is of high clinical importance. Previously, we have demonstrated that choice of a second-line therapy, in particular early in the disease course, has the capacity to prevent or delay development of secondary progressive MS form (Brown *et al*., 2019) and mitigate the evolution of neurological disability (He *et al*., 2018). An objective guidance for treatment of potentially aggressive disease at early disease stages in a condition whose management revolves around effective prevention disability driven by episodes of CNS inflammation, is an area of unmet need. The Bayesian approach used in this study allows us not only to quantify, within a year from disease onset, the probability of developing significant restriction of gait within 10 years in individual patients, but also to quantify the uncertainty across competing models – a feature that cannot be assessed within frequentist frameworks (Raftery *et al*., 1997). Reassuringly, the direction and the magnitude of the associations identified within the Bayesian and the frequentist frameworks are almost identical.

Many of these predictors share a significant proportion of variance. Our statistical approach of combining all predictors into a single analysis enables us to select only those predictors that add uniquely to the prediction. This redundancy allows us to focus the resulting predictive algorithm on a small number of accessible predictors that are able to the essence of the prediction similarly as the full set of available variables (see Table 4 and Figure 3).

**Figure 3.**
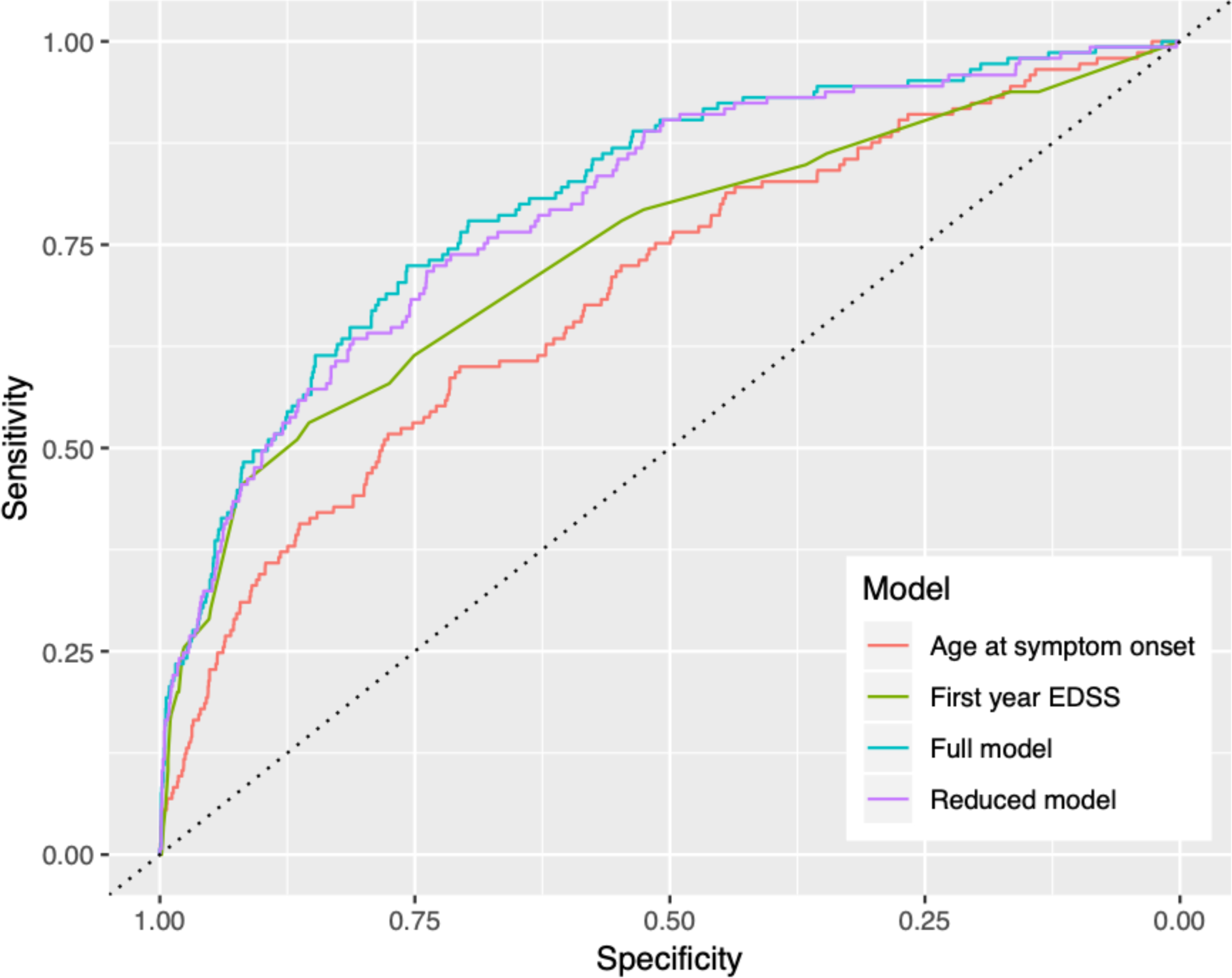
Full model = Bayesian model averaging analysis containing all predictors weighted by model fit. Reduced model = the model that contained only age at symptom onset, first year EDSS, and presence or pyramidal signs. Age at symptom onset = only the continuous predictor of age at symptom onset. First year EDSS = only the continuous predictor of median EDSS in the first year.

To the best of our knowledge, this is the largest study conducted to identify early clinical markers of aggressive MS. It enables direct translation of its results into clinical application. We employed a strict set of inclusion criteria, which required all patients to be observed within 12-months of symptom onset for a minimum of 10 years. This ensured that out control cases were truly negative in terms of aggressive MS. By requiring patients to meet criteria for aggressive MS to at least 6 months and until the end of follow-up, we ensured that the risk of false positives (i.e., patients that met aggressive disease criteria but then reverted to a negative status) was minimised. Our statistical approach was also a significant strength. Bayesian model averaging (BMA) has a long history in the statistical literature but has not been applied extensively in MS research. By averaging the results over all possible models, BMA minimises model selection bias (Raftery *et al*., 1997). This approach advances the approaches used previously to predict the course of disability. In our approach, we have not relied on a single, arbitrarily selected model, but have reported the result reflects the entire model space and combines possible statistical models.

External validity of its results is imperative for any predictive algorithm (Copas, 1983; Harrell *et al*., 1996) We therefore used an independent, population-based cohort - the Swedish MS registry, captures approximately 80% of the Swedish MS population - to validate our findings (Hillert and Stawiarz, 2015). This validation analysis confirmed that the overall model derived from the BMA analysis provided an accurate prediction in an independent MS cohort, and is thus generalisable to the prevalent MS population. At the level of the individual predictors contributing to the model, not all variables could be replicated directly. The definition of ‘motor’ signs in the Swedish MS registry differs from the definition used by MSBase. Specifically, MSBase uses the pyramidal functional system score based on EDSS(Kurtzke, 1983) while the Swedish MS registry reports motor symptoms as a separate entity. This difference is further supported by the observation of the relatively lower frequency of motor signs in the validation cohort compared to pyramidal signs in the discovery cohort. Further limitations are represented by the relative scarcity of formally reported MRI data and CSF information. However, we have conducted secondary analyses in subcohorts in whom this information was available; these analyses did not suggest that qualitatively reported MRI and CSF oligoclonal bands during the first year contribute meaningfully to the prediction of aggressive disease. Finally, the data utilised originate from observational registries based on clinical practice and are subjects to multiple sources of error and confounding. In order to mitigate these influences, we have applied an objective data quality assessment (Kalincik *et al*., 2017) and have constructed large, inclusive multivariable models that have contributed to the final model through the BMA framework.

In summary, our findings suggest that older age at symptom onset, greater disability in the first year since symptom onset, and the presence of pyramidal signs indicate a higher risk of developing aggressive disease. Importantly, the absence of any of these signs is associated with a much lower risk of aggressive disease compared to the general MS population. For ease of clinical implementation, these criteria can be defined as: median EDSS ≥ 3, any pyramidal signs on examination in the first year, and age > 35 at symptom onset. An important direction for future research will be to determine whether aggressive MS can be prevented by the use of aggressive treatment strategies in patients who are predicted to develop aggressive disease.

## Data Availability

Data were obtained from the international MSBase cohort study. Information regarding data availability can be obtained at https://www.msbase.org/.

### Appendix A Analysis of MRI and OCB data

**Table S1.** Separate BMA analyses for the subset of participants with MRI (*n*=359) and OCB *(n=*1169) data. All predictors of the full model were included.

| *Variable* | *B* | *SE* | *95% CI* | *Exp(B)* | *P (B ≠ 0 \| Y)* |
| --- | --- | --- | --- | --- | --- |
| *MRI* |  |  |  |  |  |
| Gd+ enhancing lesion | -0.02 | 0.16 | -0.02, 0.02 | 0.98 | 0.03 |
| Infratentorial lesion | -0.02 | 0.15 | -0.03, 0.02 | 0.98 | 0.04 |
| Juxtatentorial lesion | -0.00 | 0.06 | -0.01, 0.01 | 1.00 | 0.02 |
| Periventricular lesion | 0.00 | 0.08 | -0.01, 0.01 | 1.00 | 0.01 |
| *OCB* |  |  |  |  |  |
| Positive OCB | 0.03 | 0.15 | -0.01, 0.34 | 1.02 | 0.07 |

**Note:** $B$ = model coefficients computed as the mean of the posterior distribution. $SE$ = standard deviation of the posterior distributions. $CI$ = 95% credible intervals. $Exp(B)$ = odds ratios of the model coefficients. $P(B \neq 0 | Y)$ = posterior inclusion probability (PIP).

### Appendix B Investigation of fixed versus random effects models

Two models were computed to investigate whether the random effect of clinical site should be included in the final model. The first model was computed with a fixed effect for site. The second model was computed with a random intercept for site. All models were computed in *brms*, a package in the *R* environment. Parameters were estimated using the *stan* sampler with 3 chains and 2,000 samples (1,000 warmup and 1,000 actual samples). Chains and relevant metrics were inspected to ensure convergence. The same predictors were used as described in the main text. Student’s *t* priors were used for the beta and SD coefficients (location = 0, degrees of freedom = 3, scale = 2.5). A Student’s *t* prior was also placed on the intercept (location = 0, degrees of freedom = 3, scale = 10).

Model fit was assessed using leave-one-out (LOO) cross validation based on the posterior likelihood. Lower values of the LOO information criteria (LOOIC) indicate better fit. LOOICs for each model are in *Table S2* below.

**Table S2.**
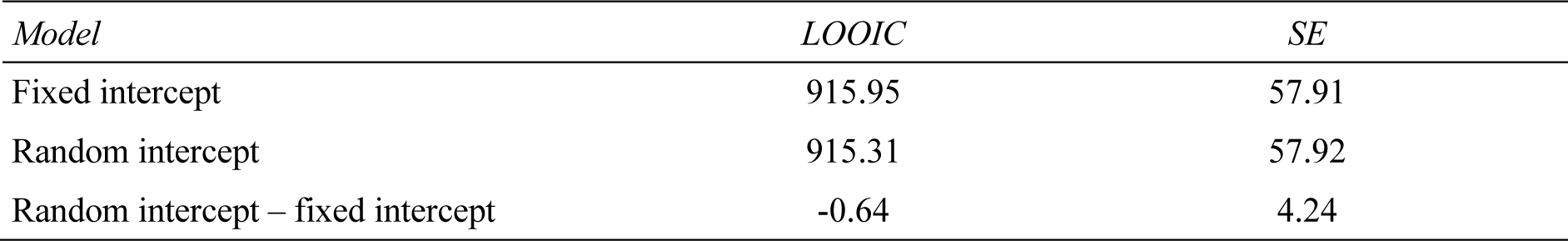
Leave-one-out information criteria (LOOIC) for fixed versus random intercept models.

As shown in *Table S2*, there was no meaningful difference between the two models. Taken together, these results do not support the added utility of including a random intercept in the model. As such, fixed intercept models were used for subsequent analyses.

### Appendix C Sensitivity analysis on baseline EDSS < 6 sub-cohort

**Table S3.** BMA analysis repeated for participants with EDSS < 6 in first year.

| *Variable* | *B* | *SD* | *95% CI* | *Exp(B)* | *P(B != 0 \| Y)* |
| --- | --- | --- | --- | --- | --- |
| *Baseline variables* |  |  |  |  |  |
| Age at symptom onset | 0.06 | 0.01 | 0.04, 0.08 | 1.06 | **1.00** |
| Male | 0.01 | 0.06 | 0.00, 0.00 | 1.01 | 0.04 |
| Disease duration at first visit | 0.00 | 0.00 | 0.00, 0.00 | 1.00 | 0.10 |
| First year EDSS | 0.58 | 0.10 | 0.39, 0.76 | 1.78 | **1.00** |
| *Events within first 12 months* |  |  |  |  |  |
| No. relapses | 0.02 | 0.07 | 0.00, 0.22 | 1.02 | 0.12 |
| No. severe relapses | 0.00 | 0.05 | 0.00, 0.00 | 1.00 | 0.03 |
| Treatment with steroids | 0.00 | 0.05 | 0.00, 0.00 | 1.00 | 0.03 |
| Hospitalisation | 0.01 | 0.08 | 0.00, 0.00 | 1.01 | 0.04 |
| Partial recovery from relapse | 0.00 | 0.04 | 0.00, 0.00 | 1.00 | 0.02 |
| Pyramidal signs | 0.37 | 0.36 | 0.00, 0.94 | 1.45 | **0.57** |
| Bowel/bladder signs | 0.03 | 0.14 | 0.00, 0.32 | 1.03 | 0.06 |
| Cerebellar signs | 0.11 | 0.25 | 0.00, 0.71 | 1.12 | 0.21 |
| *Treatment* |  |  |  |  |  |
| % time on first-line (10y) | -0.01 | 0.06 | -0.02, 0.01 | 0.99 | 0.02 |
| % time on second-line (10y) | 2.24 | 0.54 | 1.29, 3.30 | 9.42 | **0.99** |
| % time on first-line (1y) | -0.02 | 0.11 | -0.03, 0.02 | 0.98 | 0.04 |
| % time on second-line (1y) | -0.04 | 0.34 | -0.05, 0.03 | 0.96 | 0.03 |

**Note:** *B* = model coefficients computed as the mean of the posterior distribution. SE = standard deviation of the posterior distributions. CI = 95% credible intervals. Exp(*B*) = odds ratios of the model coefficients. *P*(*B* != 0 | *Y*) = posterior inclusion probability (PIP).

**Table S4.** Frequentist analysis repeated for participants with EDSS < 6 in first year.

| *Variable* | *B* | *SE* | *95% CIs* | *Exp(B)* | *Sig.* |
| --- | --- | --- | --- | --- | --- |
| *Baseline variables* |  |  |  |  |  |
| Age at symptom onset | 0.06 | 0.01 | 0.04, 0.08 | 1.06 | **<0.001** |
| Male | 0.27 | 0.21 | -0.15, 0.68 | 1.31 | 0.206 |
| Disease duration at first visit | 0.00 | 0.00 | 0.00, 0.00 | 1.00 | 0.181 |
| First year EDSS | 0.48 | 0.11 | 0.26, 0.7 | 1.62 | **<0.001** |
| *Events within first 12 months* |  |  |  |  |  |
| No. relapses | 0.17 | 0.12 | -0.08, 0.41 | 1.18 | 0.182 |
| No. serious relapses | -0.31 | 0.24 | -0.78, 0.16 | 0.73 | 0.192 |
| Treatment with steroids | 0.04 | 0.28 | -0.51, 0.58 | 1.04 | 0.895 |
| Hospitalisation | 0.22 | 0.35 | -0.46, 0.90 | 1.25 | 0.525 |
| Partial recovery from relapse | -0.04 | 0.34 | -0.71, 0.62 | 0.96 | 0.898 |
| Pyramidal signs | 0.55 | 0.24 | 0.08, 1.02 | 1.73 | **0.021** |
| Bowel/bladder signs | 0.37 | 0.31 | -0.24, 0.98 | 1.45 | 0.235 |
| Cerebellar signs | 0.39 | 0.24 | -0.07, 0.85 | 1.48 | 0.098 |
| *Treatment* |  |  |  |  |  |
| % time on first-line (10y) | -0.26 | 0.32 | -0.89, 0.37 | 0.77 | 0.414 |
| % time on second-line (10y) | 2.23 | 0.59 | 1.06, 3.39 | 9.27 | **<0.001** |
| % time on first-line (1y) | -0.25 | 0.45 | -1.13, 0.62 | 0.78 | 0.571 |
| % time on second-line (1y) | -1.31 | 1.65 | -4.55, 1.93 | 0.27 | 0.429 |

**Note:** *B* = model coefficients. *SE* = robust standard errors computed via heteroskedasticity-consistent sandwich estimation. *CI* = 95% confidence intervals computed using robust *SEs*. *Exp(B)* = odds ratios for the model coefficients. *Sig* = significance values were computed using the robust *SEs*. Bold significance values indicate $p < .05$ . Assumption checks regarding multicollinearity, influential cases, autocorrelation, dispersion, and residual distribution were satisfied.

### Appendix D Cross-validation of predictive model

**Figure S1.**
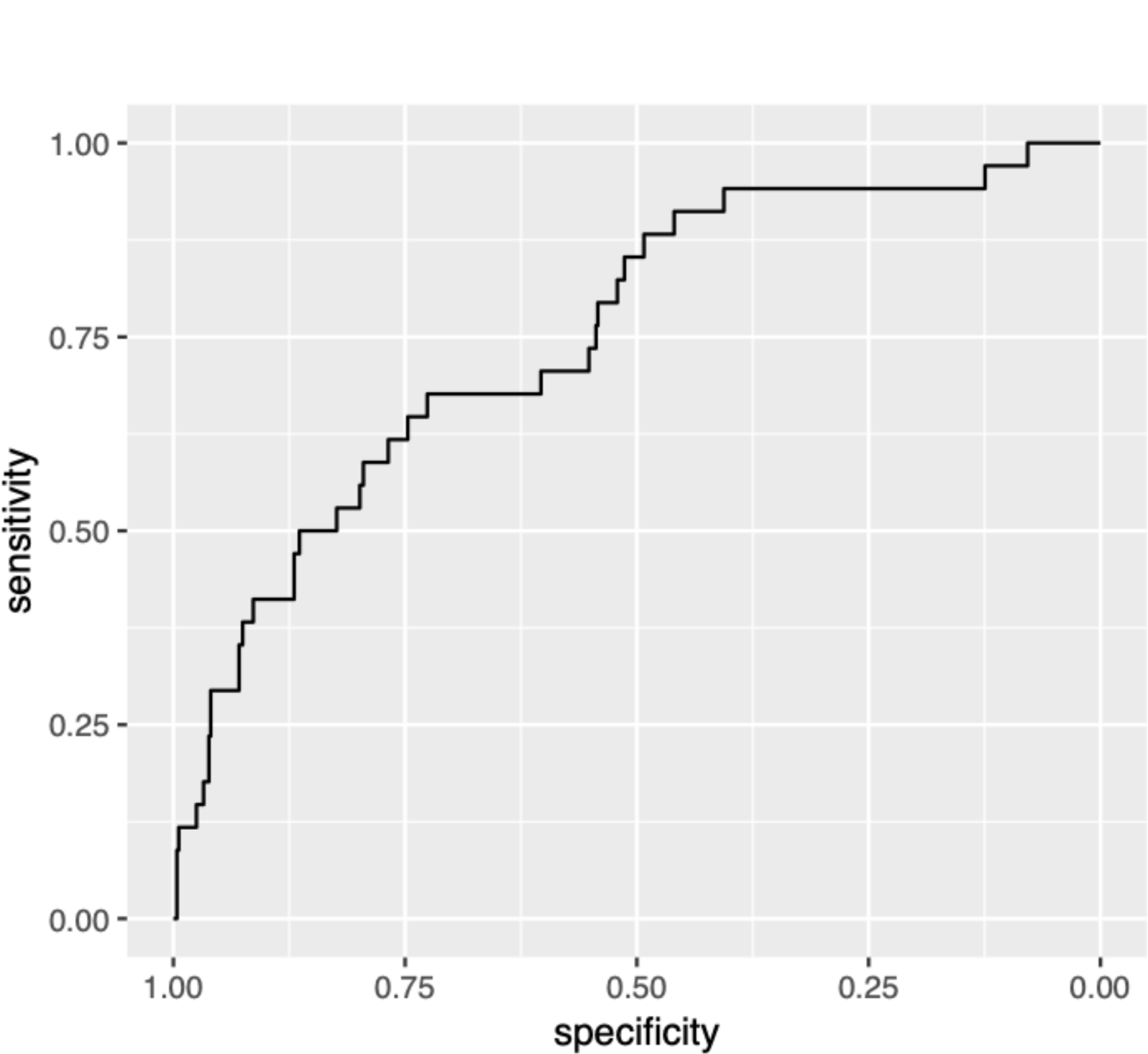
Receiver operating characteristic (ROC) curve for the validation of the three-predictor model in the Swedish MS Registry cohort.

## Notes

Ali Manouchehrinia is supported by Margaretha af Ugglas foundation. He has received consultancy fees from Biogen and speaker honoraria and/or travel expenses from Biogen and ECTRIMS.

### Competing Interest Statement

Charles Malpas: nothing to disclose.
Ali Manouchehrinia is supported by Margaretha af Ugglas foundation. He has received consultancy fees from Biogen and speaker honoraria and/or travel expenses from Biogen and ECTRIMS.
Sifat Sharmin: nothing to disclose
Dana Horakova received speaker honoraria and consulting fees from Biogen, Merck, Teva and Novartis, as well as support for research activities from Biogen and research grants from Charles University in Prague [PRVOUK-P26/LF1/4], Czech Minsitry of Education [PROGRES Q27/LF1] and Czech Ministry of Health [NT13237-4/2012].
Eva Kubala Havrdova received speaker honoraria and consultant fees from Actelion, Biogen, Celgene, Merck, Novartis, Roche, Sanofi and Teva, and support for research activities from Czech Ministry of Education [project Progres Q27/LF1].
Maria Trojano received speaker honoraria from Biogen-Idec, Bayer-Schering, Sanofi Aventis, Merck, Teva, Novartis and Almirall; has received research grants for her Institution from Biogen-Idec, Merck, and Novartis.
Guillermo Izquierdo received speaking honoraria from Biogen, Novartis, Sanofi, Merck, Roche, Almirall and Teva.
Sara Eichau received speaker honoraria and consultant fees from Biogen, Merck,
Novartis, Almirall, Roche, Sanofi and Teva.
Roberto Bergamaschi received speaker honoraria from Bayer Schering, Biogen, Genzyme, Merck, Novartis, Sanofi-Aventis, Teva; research grants from Bayer Schering, Biogen, Merck, Novartis, Sanofi-Aventis, Teva; congress and travel/accommodation expense compensations by Almirall, Bayer Schering, Biogen, Genzyme, Merck, Novartis, Sanofi-Aventis, Teva.
Patrizia Sola served on scientific advisory boards for Biogen Idec and TEVA, she has received funding for travel and speaker honoraria from Biogen Idec, Merck, Teva, Sanofi Genzyme, Novartis and Bayer and research grants for her Institution from Bayer, Biogen, Merck, Novartis, Sanofi, Teva.
Diana Ferraro received travel grants and/or speaker honoraria from Merck, TEVA,?Novartis, Biogen and Sanofi-Genzyme.
Alessandra Lugaresi is a Bayer, Biogen, Genzyme, Merck Advisory Board Member. She received travel grants and honoraria from Roche, Bayer, Biogen, Merck, Novartis, Sanofi, Teva and Fondazione Italiana Sclerosi Multipla (FISM). Her institution received research grants from Bayer, Biogen, Merck, Novartis, Sanofi, Teva and Fondazione Italiana Sclerosi Multipla (FISM).
Alexandre Prat did not declare any competing interests.
Marc Girard received consulting fees from Teva Canada Innovation, Biogen, Novartis and Genzyme Sanofi; lecture payments from Teva Canada Innovation, Novartis and EMD.? He has also received a research grant from Canadian Institutes of Health Research.
Pierre Duquette served on editorial boards and has been supported to attend meetings by EMD, Biogen, Novartis, Genzyme, and TEVA Neuroscience. He holds grants from the CIHR and the MS Society of Canada and has received funding for investigator-initiated trials from Biogen, Novartis, and Genzyme.
Pierre Grammond is a Merck, Novartis, Teva-neuroscience, Biogen and Genzyme advisory board member, consultant for Merck, received payments for lectures by Merck, Teva-Neuroscience and Canadian Multiple sclerosis society, and received grants for travel from Teva-Neuroscience and Novartis.
Francois Grand'Maison received honoraria or research funding from Biogen, Genzyme, Novartis, Teva Neurosciences, Mitsubishi and ONO Pharmaceuticals.
Serkan Ozakbas did not declare any competing interests.
Vincent Van Pesch received travel grants from Biogen, Bayer Schering, Genzyme, Merck, Teva and Novartis Pharma. His institution receives honoraria for consultancy and lectures from Biogen, Bayer Schering, Genzyme, Merck, Roche, Teva and Novartis Pharma as well as research grants from Novartis Pharma and Bayer Schering.
Franco Granella received research grant from Biogen, served on scientific advisory boards for Biogen, Novartis, Merck, and Sanofi-Aventis and received funding for travel and speaker honoraria from Biogen, Merck, Sanofi-Aventis, and Almirall.
Raymond Hupperts received honoraria as consultant on scientific advisory boards from Merck, Biogen, Genzyme-Sanofi and Teva, research funding from Merck and Biogen, and speaker honoraria from Sanofi-Genzyme and Novartis.
Eugenio Pucci served on scientific advisory boards for Merck, Genzyme and Biogen; he has received honoraria and travel grants from Sanofi Aventis, Novartis, Biogen, Merck, Genzyme and Teva; he has received travel grants and equipment from "Associazione Marchigiana Sclerosi Multipla e altre malattie neurologiche".
Tomas Kalincik served on scientific advisory boards for Roche, Genzyme-Sanofi, Novartis, Merck and Biogen, steering committee for Brain Atrophy Initiative by Genzyme, received conference travel support and/or speaker honoraria from WebMD Global, Novartis, Biogen, Genzyme-Sanofi, Teva, BioCSL and Merck and received research support from Biogen.
Cavit Boz received conference travel support from Biogen, Novartis, Bayer-Schering, Merck and Teva; has participated in clinical trials by Sanofi Aventis, Roche and Novartis.
Gerardo Iuliano had travel/accommodations/meeting expenses funded by Bayer Schering, Biogen, Merck, Novartis, Sanofi Aventis, and Teva.
Youssef Sidhom did not declare any competing interests.
Riadh Gouider did not declare any competing interests.
Daniele Spitaleri received honoraria as a consultant on scientific advisory boards by Bayer-Schering, Novartis and Sanofi-Aventis and compensation for travel from Novartis, Biogen, Sanofi Aventis, Teva and Merck.
Helmut Butzkueven served on scientific advisory boards for Biogen, Novartis and Sanofi-Aventis and has received conference travel support from Novartis, Biogen and Sanofi Aventis. He serves on steering committees for trials conducted by Biogen and Novartis, and has received research support from Merck, Novartis and Biogen.
Aysun Soysal did not declare any competing interests.
Thor Petersen received funding or speaker honoraria from Biogen, Merck, Novartis, Bayer Schering, Sanofi-Aventis, Roche, and Genzyme.
Freek Verheul is an advisory board member for Teva Biogen Merck and Novartis.
Rana Karabudak did not declare any competing interests.
Recai Turkoglu did not declare any competing interests.
Cristina Ramo-Tello received research funding, compensation for travel or speaker honoraria from Biogen, Novartis, Genzyme and Almirall.
Murat Terzi received travel grants from Novartis, Bayer-Schering, Merck and Teva; has participated in clinical trials by Sanofi Aventis, Roche and Novartis.
Edgardo Cristiano received honoraria as consultant on scientific advisory boards by Biogen, Bayer-Schering, Merck, Genzyme and Novartis; has participated in clinical trials/other research projects by Merck, Roche and Novartis.
Mark Slee has participated in, but not received honoraria for, advisory board activity for Biogen, Merck, Bayer Schering, Sanofi Aventis and Novartis.
Pamela McCombe did not declare any competing interests.
Richard Macdonell did not declare any competing interests.
Yara Fragoso received honoraria as a consultant on scientific advisory boards by Novartis, Teva, Roche and Sanofi-Aventis and compensation for travel from Novartis, Biogen, Sanofi Aventis, Teva, Roche and Merck.
Javier Olascoaga serves on scientific advisory boards for Biogen, Novartis, Sanofi and Roche; has received speaker honoraria from Almirall, Biogen, Bayer, Sanofi, Merck, Novartis and Roche and research grants from Biogen, Merck, Novartis and Teva.
Ayse Altintas received personal fees and speaker honoraria from Teva, Merck, Novartis, Sanofi-Genzyme; received travel and registration grants from Merck, Roche, Sanofi-Genzyme.
Tomas Olsson has received honoraria for lectures/advisory boards and/or unrestricted MS research grants from Biogen, Novartis, Sanofi, Merck and Roche
Tomas Kalincik served on scientific advisory boards for Roche, Genzyme-Sanofi, Novartis, Merck and Biogen, steering committee for Brain Atrophy Initiative by Genzyme, received conference travel support and/or speaker honoraria from WebMD Global, Novartis, Biogen, Genzyme-Sanofi, Teva, BioCSL and Merck and received research support from Biogen.

### Funding Statement

This study was financially supported by the National Health and Medical Research Council of Australia (1129189, 1140766, 1080518). The MSBase Foundation is a not-for-profit organisation that receives support from Roche, Merck, Biogen, Novartis, Bayer Schering, Sanofi Genzyme and Teva. The study was conducted separately and apart from the guidance of the sponsors.

### Author Declarations

All relevant ethical guidelines have been followed and any necessary IRB and/or ethics committee approvals have been obtained.

Any clinical trials involved have been registered with an ICMJE-approved registry such as ClinicalTrials.gov and the trial ID is included in the manuscript.

